# A proteome atlas of structural variation and VNTR effects on complex traits and diseases

**DOI:** 10.64898/2026.09.26.26364064

**Authors:** Peixiong Yuan, Wei-Yang Bai, Junren Hou, Jian Yang

**Affiliations:** New Cornerstone Science Laboratory, School of Life Sciences, Westlake University, Hangzhou, Zhejiang 310024, China; Westlake Laboratory of Life Sciences and Biomedicine, Hangzhou, Zhejiang 310024, China

## Abstract

Structural variants (SVs), including variable number tandem repeats (VNTRs), are a major source of human genetic variation, but their impact on the proteome remains poorly characterized. Using a long-read assembly-based reference panel, we imputed 54,578 general SVs and 15,826 VNTRs in 54,306 UK Biobank participants to interrogate their effects on the abundances of 2,923 plasma proteins. SVs and VNTRs together explained ∼8.0% of variant-based heritability for protein abundance, with 123 proteins driven predominantly (>80%) by SVs. We identified 8,065 independent SV-protein and 4,101 VNTR-protein associations (with 601 highly unlikely to be driven by small genetic variants) that frequently perturb active regulatory elements, topologically associating domain boundaries, and post-transcriptional mechanisms. Integrative analyses with gene expression and complex trait SV association data identified 1,353 protein-trait association pairs, such as a highly pleiotropic insertion in *MAN1A2* associated with 86 distinct phenotypes and a 3’ UTR deletion in *SULT2A1* linked to gallstone disease, illustrating plausible mechanisms whereby SV-induced regulatory perturbations mediate disease susceptibility. This SV-protein atlas bridges a critical gap between large-scale genomic alterations and proteomic variation, providing novel mechanistic insights into human disease biology and therapeutic discovery.

## Introduction

Structural variants (SVs), including deletions, insertions, inversions, and other complex forms, are large-scale genomic alterations ranging from 50 bp to several megabases. Compared to small genetic variants (SGVs), such as single-nucleotide polymorphisms (SNPs) and short insertions and deletions (indels), SVs are longer and thus more likely to disrupt protein-coding sequences or regulatory elements, making them a major source of human genetic diversity^1,2^. Accumulating evidence has highlighted the substantial contributions of SVs to phenotypic variation and disease susceptibility^3–5^. Many of these effects are mediated by proteins, the primary functional outputs of gene regulation and key determinants of cellular pathways, biomarkers, and therapeutic targets^6–8^. Despite this growing recognition, the impact of SVs on protein abundances remains far less explored than that of SGVs.

To date, protein quantitative trait locus (pQTL) studies have largely focused on SGVs, leveraging SNP arrays^9^, short-read whole-exome sequencing^10^, or short-read whole-genome sequencing (WGS)^11^. While recent pioneering efforts have begun to extend these analyses to SVs, such as utilizing short-read WGS to map SV-pQTLs for 2,907 proteins^11^, systematic investigations of SV-associated pQTLs (SV-pQTLs) remain limited, primarily due to challenges in accurate SV detection and genotyping at scale. This is because although short-read sequencing enables genome-wide SV discovery, its limited sensitivity and breakpoint resolution, particularly in repetitive or structurally complex regions, restrict the number of SVs that can be reliably identified^12^. For example, SV callsets derived from short-read WGS in large biobank cohorts typically capture 9,000-13,000 SVs per individual, substantially fewer than the ∼26,000 SVs detectable using high-accuracy long-read sequencing^13,14^. While long-read technologies provide a more comprehensive view of SV landscapes, their current cost remains prohibitive for population-scale studies^12^. As a result, the lack of high-quality SV genotypes across large cohorts has been a major bottleneck for comprehensive SV-pQTL discovery.

Imputation of SVs using high-quality reference panels offers a scalable and cost-effective solution to this challenge, enabling genome-wide SV profiling in large biobanks based on existing SNP array data^4,14^. Previous attempts to perform protein association analyses using imputed SVs (covering 1,463 proteins, approximately half of the currently available plasma proteins in the UKB) relied on reference panels constructed from moderate-coverage Oxford Nanopore Technology (ONT) sequencing, which resulted in limited imputation accuracy and a restricted number of well-imputed variants^4^. These limitations underscore the critical importance of sequencing accuracy and reference panel quality for reliable SV imputation. Recently, we developed a comprehensive, multi-ancestry SV imputation panel based on 482 haplotype-resolved assemblies from 241 individuals, generated using high-coverage PacBio HiFi long-read WGS. This resource captures 171,233 high-quality general SVs and 18,360 variable number tandem repeats (VNTRs), providing broad genomic coverage and high imputation accuracy^5^.

In this study, we imputed SVs and VNTRs into the UKB and systematically evaluated their contributions to the abundances of 2,923 circulating plasma proteins measured in 54,306 participants. We mapped SV- and VNTR-associated pQTLs to build an SV-protein atlas. Integration of this atlas with SV-based expression quantitative trait locus (eQTL) and genome-wide association study (GWAS) data enabled us to assess the functional relevance of these associations and to gain insights into the roles of SVs and VNTRs in complex traits and disease biology (Fig. 1).

**Figure 1.**
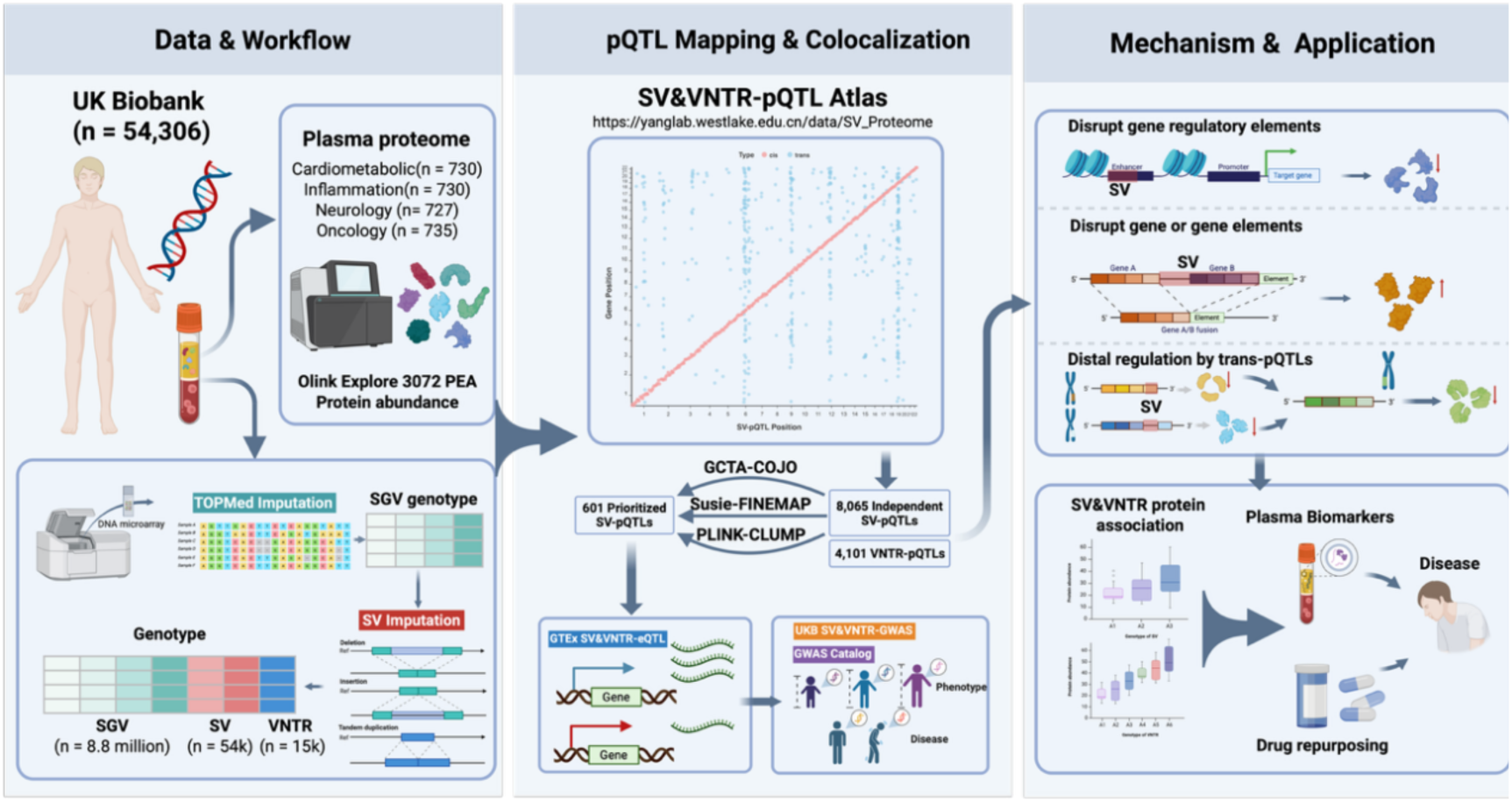
Study overview. Plasma proteomic data from 54,306 UK Biobank participants were integrated with genotypes of small genetic variants (SGVs), structural variants (SVs), and VNTRs, utilizing long-read assembly-based imputation of SVs and VNTRs. Genome-wide pQTL mapping identified SV- and VNTR-pQTLs, followed by prioritization using PLINK clumping, GCTA-COJO, and SuSiE fine-mapping, and colocalization with GTEx eQTLs, and UK Biobank GWAS traits and the GWAS Catalog. Representative mechanisms show that SVs can affect protein abundance through the disruption of regulatory elements, gene-body variation, or distal *trans* effects. These findings provide a resource for biomarker discovery, disease interpretation, and drug repurposing.

## Results

### Heritability contributions of SVs and VNTRs to plasma proteins

We analyzed antibody-based proteomic measurements (Olink Explore) for 2,923 plasma proteins from 54,306 UKB participants of European ancestry via the UKB Pharma Proteomics Project (UKB-PPP)^9^. These proteins were grouped into four panels: cardiometabolic (*m* = 730), inflammation (*m* = 730), neurology (*m* = 727), and oncology (*m* = 735). We imputed SVs into UKB and retained those meeting the following quality control criteria: minor allele frequency (MAF) ≥ 0.01, imputation INFO score ≥ 0.3, and no deviation from Hardy-Weinberg equilibrium (*P* > 1 × 10^−6^). For VNTRs, only loci with at least two distinct non-reference alleles were included. After filtering, the final dataset comprised 54,578 common SVs and 15,826 VNTRs for downstream analyses (Fig. 2a,b, Supplementary Fig. 1). We also included ∼8.8 million TOPMed-imputed SGVs with MAF ≥ 0.01 available from the UKB^15,16^.

**Figure 2.**
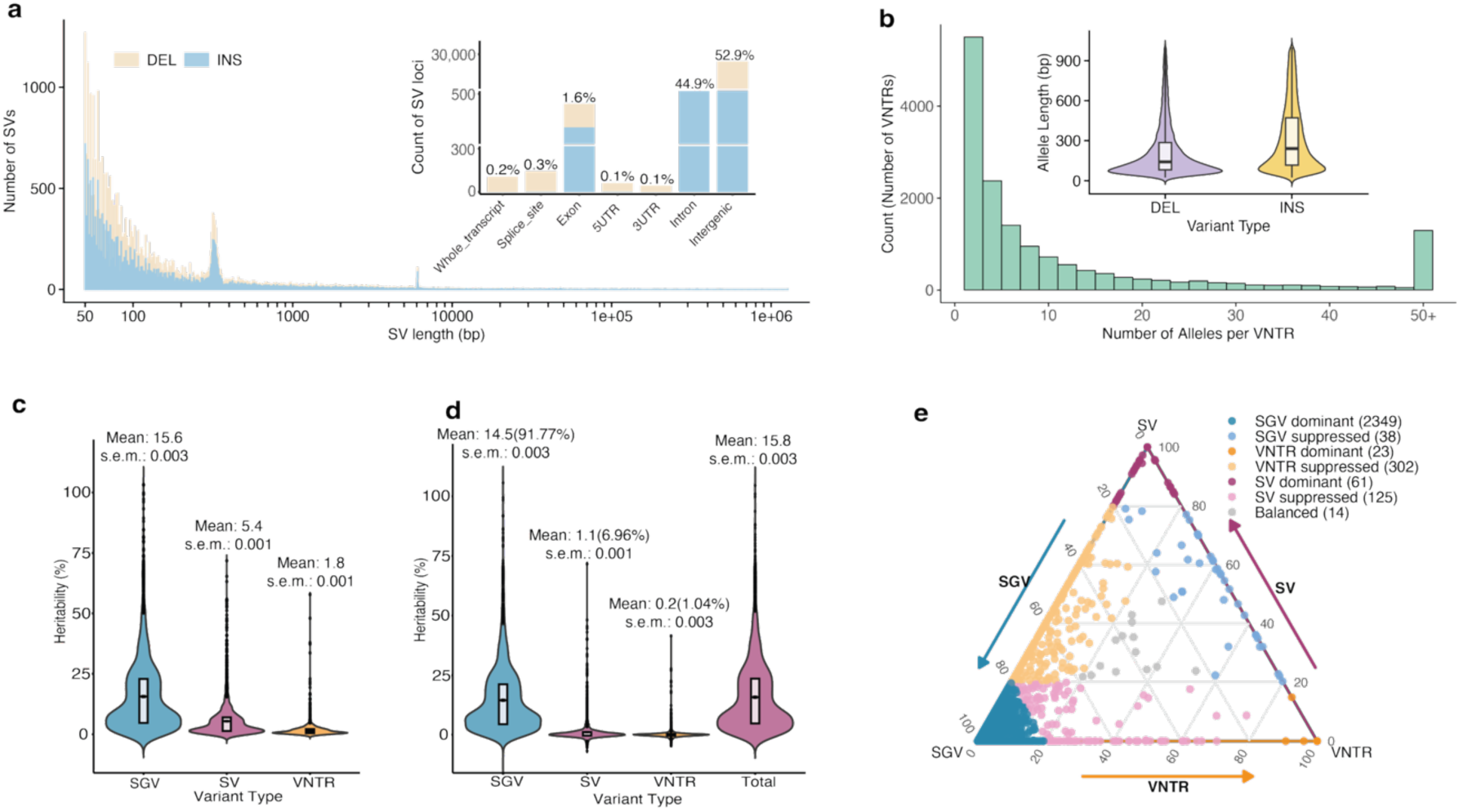
Contribution of SVs and VNTRs to the variant-based heritability of protein abundance. a) Distribution of SV lengths. The peaks indicate *Alu*, SVA, and LINE elements. The genomic context of SVs is summarized by their overlap with different gene features. b) Distributions of VNTR allele copy numbers and motif lengths. c) Distribution of heritability estimates obtained by fitting separate GRMs computed for each variant class, with heritability estimated independently for each class. d) Distribution of heritability estimates obtained from a joint model that includes GRMs from multiple variant classes simultaneously. e) Distribution of the proportion of heritability explained by each variant class across 2,922 proteins. ‘Dominant’ indicates that a given variant class accounts for >80% of the heritability; ‘Suppressed’ indicates a contribution ≤20%; and ‘Balanced’ denotes proteins for which the contributions of the three variant classes are relatively even and do not meet the above thresholds.

For each of the 2,922 proteins passing quality control (Methods), we used GCTA-GREML^17,18^ to estimate the proportion of phenotypic variance explained by all imputed SGVs, SVs, and VNTRs (referred to as variant-based heritability), denoted by 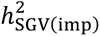, 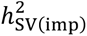 and 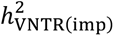, respectively. We evaluated both a joint model, in which the three variant classes were fitted simultaneously as three random-effect components, and marginal models, in which each class was fitted separately. The proportion of variance explained by all imputed variants analyzed in this study is denoted by 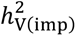. In the marginal analyses, SGVs explained an average of 15.6% of the phenotypic variance across proteins (s. e. m. = 0.003; Fig. 2c,d; Supplementary Tables 1-2).

We observed a specific subset of proteins with a high proportion of 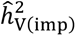 attributable to SVs. Notably, 123 proteins exhibited 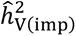 almost entirely dominated by SVs and VNTRs (≥80% of 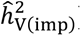, whereas 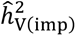 for the majority of proteins remained predominantly explained by SGVs (Fig. 2e). Two striking examples illustrate cases in which SVs play a dominant role in shaping protein abundance. The first is a well-imputed insertion (INFO score = 0.89) of 192 bp (denoted 14q11.2-INS-192bp; chr14:25,505,261), located in the 5’ untranslated region (UTR) of the protein-coding gene *RNASE10*. Incorporation of SVs into the variant-based heritability model resulted in a marked increase in 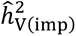 of RNASE10 protein abundance, from 0.08 (*s*. *e*. = 0.011) to 0.51 (*s*. *e*. = 0.010).

Approximately 88% of 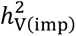 for the abundance of RNASE10 was attributable to this single insertion (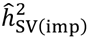 = 0.45; s. e. = 0.009). The second example is a large deletion spanning 16.4 kbp (denoted 19q13.41-DEL-16.4kbp; chr19:51,630,515-51,646,938) that completely removes the *SIGLEC14* gene. This deletion has previously been reported to generate a fusion allele placing *SIGLEC5* under the control of the *SIGLEC14* promoter^19^, leading to reduced *SIGLEC5* expression in whole blood. In our analysis, the 19q13.41-DEL-16.4kbp deletion accounted for approximately 85% of 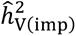 for the abundance of SIGLEC5 (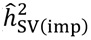 = 0.47; s. e. = 0.011).

### Genome-wide association analysis of SVs with 2,922 plasma proteins in UKB

We performed genome-wide association analyses for SVs and SGVs in 53,013 individuals using GCTA-fastGWA^20,21^, with sample relatedness accounted for by a sparse genetic relationship matrix. We identified 8,065 significant, independent SV-pQTLs at an experiment-wise significance threshold of *P*_SV−pQTL_ < 1.7 × 10^−11^ (5 × 10^−8^ ∕ 2,922), including 4,030 *cis* associations (∼50%; variants located within 1 Mb of the gene encoding the target protein) and 4,035 *trans* associations (∼50%). These signals involved 4,319 distinct SVs and 1,816 proteins, corresponding to 62% of all assayed proteins (Fig. 3a, Supplementary Table 3). All association results, along with detailed annotations, are available at our interactive SV-pQTL portal (https://yanglab.westlake.edu.cn/data/sv-pqtl).

**Figure 3.**
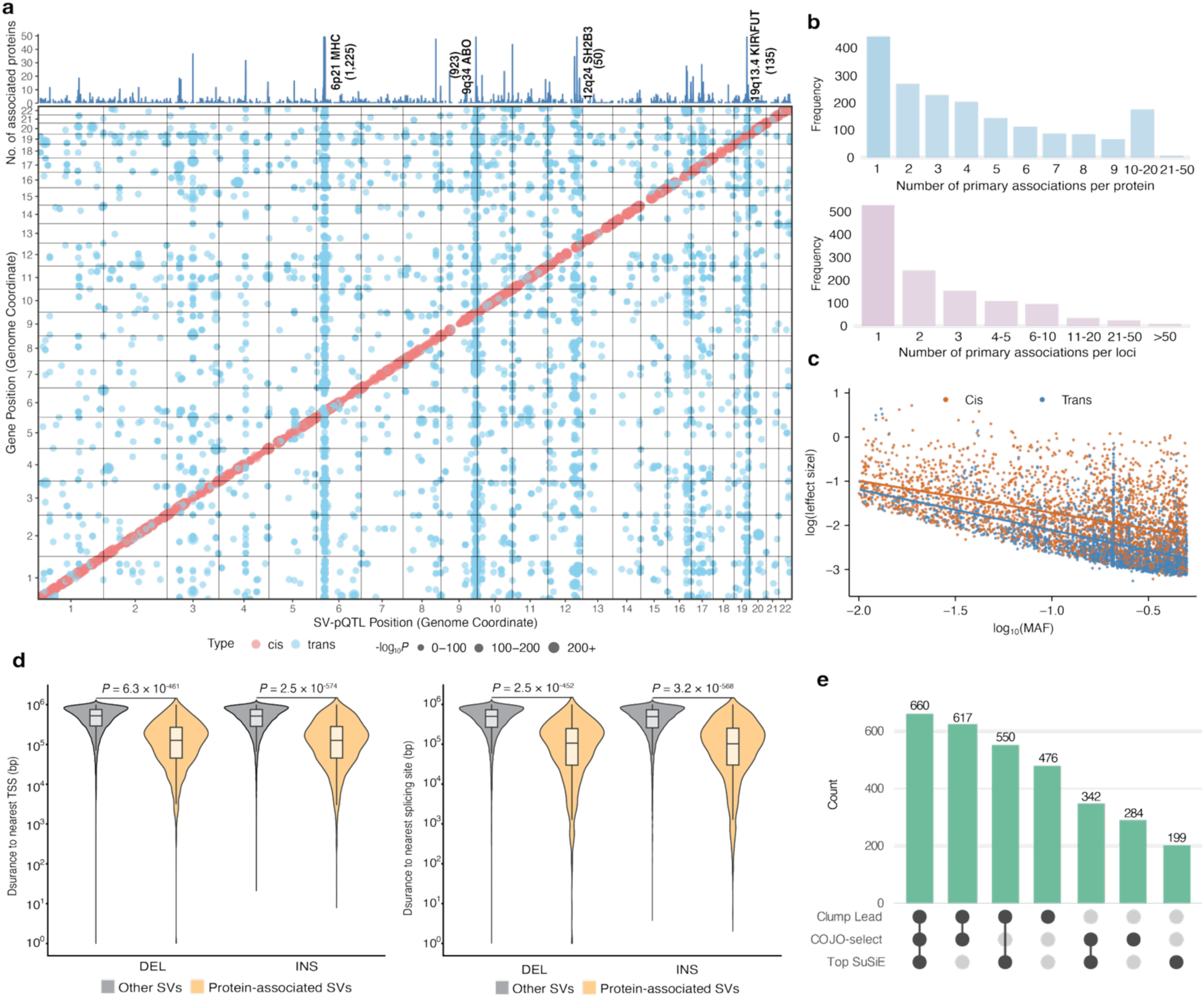
Descriptive summary of genome-wide SV-protein association analysis. a) Genome-wide SV-protein association results. The bottom panel shows the genomic distribution of SV-pQTLs across the genome, with SV genomic position on the x-axis and gene position on the y-axis; point size indicates the strength of association. The top panel summarizes the number of proteins with significant associations in each genomic region, with several hotspot loci and their corresponding counts annotated, including the MHC region, the *ABO* locus (glycosylation), the *SH2B3* locus (a key regulatory adaptor protein in hematopoietic cells), the KIR region (regulating NK-cell cytotoxicity and cytokine secretion), and the *FUT* locus (glycosyltransferase). b) Top: frequency distribution of the number of significant variant associations per protein. Bottom: frequency distribution of the number of proteins associated with each locus. c) Changes in effect size with MAF for *cis-*pQTLs and *trans-* pQTLs. d) Left: distance to the nearest splice site (SS) for significant, independent *cis*-pQTL deletions (DEL) and insertions (INS) compared with non-significant, independent variants within the same *cis* window (TSS ± 1 Mb). Right: distance to the nearest transcript start site (TSS) for significant, independent *cis*-pQTL DELs and INSs compared with non-significant, independent variants. *P* values were calculated using two-sided Wilcoxon rank-sum tests. e) Union of SV-pQTLs prioritized by different approaches: lead SV-pQTLs identified by LD clumping, SV-pQTLs remaining significant after COJO conditioning on nearby SGVs, and top-PIP SV-pQTLs identified by SuSiE fine-mapping, shown as the distribution of counts across methods.

Across proteins, the number of SV-pQTLs per protein ranged from 1 to 34 (mean = 4.4), with 75.7% of proteins (1,375 out of 2,922) harboring at least two SV-pQTL signals. From the variant perspective, 1,111 SV loci (25.7%) were associated with multiple proteins (Fig. 3b), indicating widespread pleiotropic effects. We next examined the relationship between pQTL effect size and MAF. For both *cis* and *trans* associations, effect sizes increased with decreasing MAF, with *cis* SV-pQTLs consistently exhibiting larger effects than *trans* SV-pQTLs across the entire MAF spectrum (Fig. 3c), consistent with an effect of negative selection as observed for SGVs in prior work^9^. We also compared the genomic locations of significant *cis*-pQTL SVs and non-significant SVs within the same *cis* window (1 Mb of the transcription start site (TSS) of the cognate gene). Both deletions and insertions with significant *cis*-pQTL associations were located considerably closer to the TSS (DEL: *P* = 6.3 × 10^−461^, INS: *P* = 2.5 × 10^−574^ , two-sided Wilcoxon rank-sum test) and to splice sites (SS) (DEL : *P* = 2.5 × 10^−452^, INS: *P* = 3.2 × 10^−568^) compared to non-significant SVs (Fig. 3d).

Previous SGV-pQTL studies using the Olink and SomaScan platforms have shown that a substantial fraction of *cis*-pQTLs can arise from epitope-binding artifacts^22^. Protein-altering variants (PAVs) located within the coding sequences or splice regions of the cognate gene can alter the binding affinity of the assay reagents, generating spurious association signals^8^. We therefore applied a pragmatic filtering strategy to our 8,065 primary SV-protein associations. Specifically, a locus was considered potentially affected by epitope-binding artifacts if it correlated with a PAV (linkage disequilibrium (LD) *r*^2^ > 0.1) and simultaneously lacked supporting evidence from *cis* SV-eQTL signals at the gene expression level. Based on these criteria, we identified 240 loci (6.0%) as possibly influenced by epitope effects (Supplementary Table 4), yielding 7,825 confident SV-protein association pairs. To further assess the robustness of our findings, we conducted sensitivity analyses following established approaches^9^. We repeated the association tests with additional adjustments for blood collection time, blood cell counts, and body mass index (BMI), factors known to potentially influence plasma protein measurements. The resulting association patterns were highly consistent with the primary analyses, indicating the robustness of the SV-pQTL signals (Supplementary Fig. 2).

To distinguish SV-pQTLs representing distinct structural regulatory mechanisms from those merely tagging SGV effects at the same locus, we applied a stringent variant prioritization framework. This was informed by our previous observation that imputed SV associations are highly unlikely (frequency < 0.6%) to be driven by SGVs if the SV satisfies at least one of three conditions: it is the lead variant at the locus, it remains genome-wide significant after conditioning on nearby SGVs, or it attains the highest posterior inclusion probability (PIP) in SuSiE fine-mapping analyses^5^. We first performed LD-based clumping of genome-wide significant SV-pQTLs and SGV-pQTLs using PLINK^23^, which identified 454 independent lead SV associations (Supplementary Table 5). Next, conditional analyses using GCTA-COJO^24^ identified 247 SVs that remained genome-wide significant after conditioning on nearby SGVs (Supplementary Table 6). Finally, fine-mapping analysis using SuSiE^25^ of pQTL loci including both SVs and SGVs identified 188 cases where the SV attained the highest PIP^25^ (Supplementary Table 7). Collectively, these complementary analyses prioritized 601 unique SV-protein associations that represent distinct structural effects rather than mere tags for linked SGVs (Fig. 3e).

### Functional enrichment analysis of SV-pQTLs

To explore the mechanisms underlying these SV-protein associations, we investigated whether SV-pQTLs are enriched within functionally active regulatory elements. Specifically, we compared genome-wide significant SV-pQTLs with non-significant SVs across multiple regulatory annotations, including topologically associating domain (TAD) boundaries^26^, RNA-binding protein (RBP) binding sites, histone modifications^27^, and chromatin-state annotations^28^. Our results suggested that SV-pQTLs were markedly enriched at TAD boundaries (Fig. 4b) and RBP binding sites (Fig. 4d), implicating both 3D chromatin architecture and post-transcriptional regulation in shaping SV effects on protein levels. Furthermore, analysis of histone modifications (Fig. 4c) and chromatin states (Fig. 4a) revealed that SV-pQTLs were preferentially located in enhancer-like states and depleted from repressive or quiescent regions, indicating a strong preference for active regulatory elements. We highlight two representative examples below that illustrate the underlying regulatory roles of SVs.

**Figure 4.**
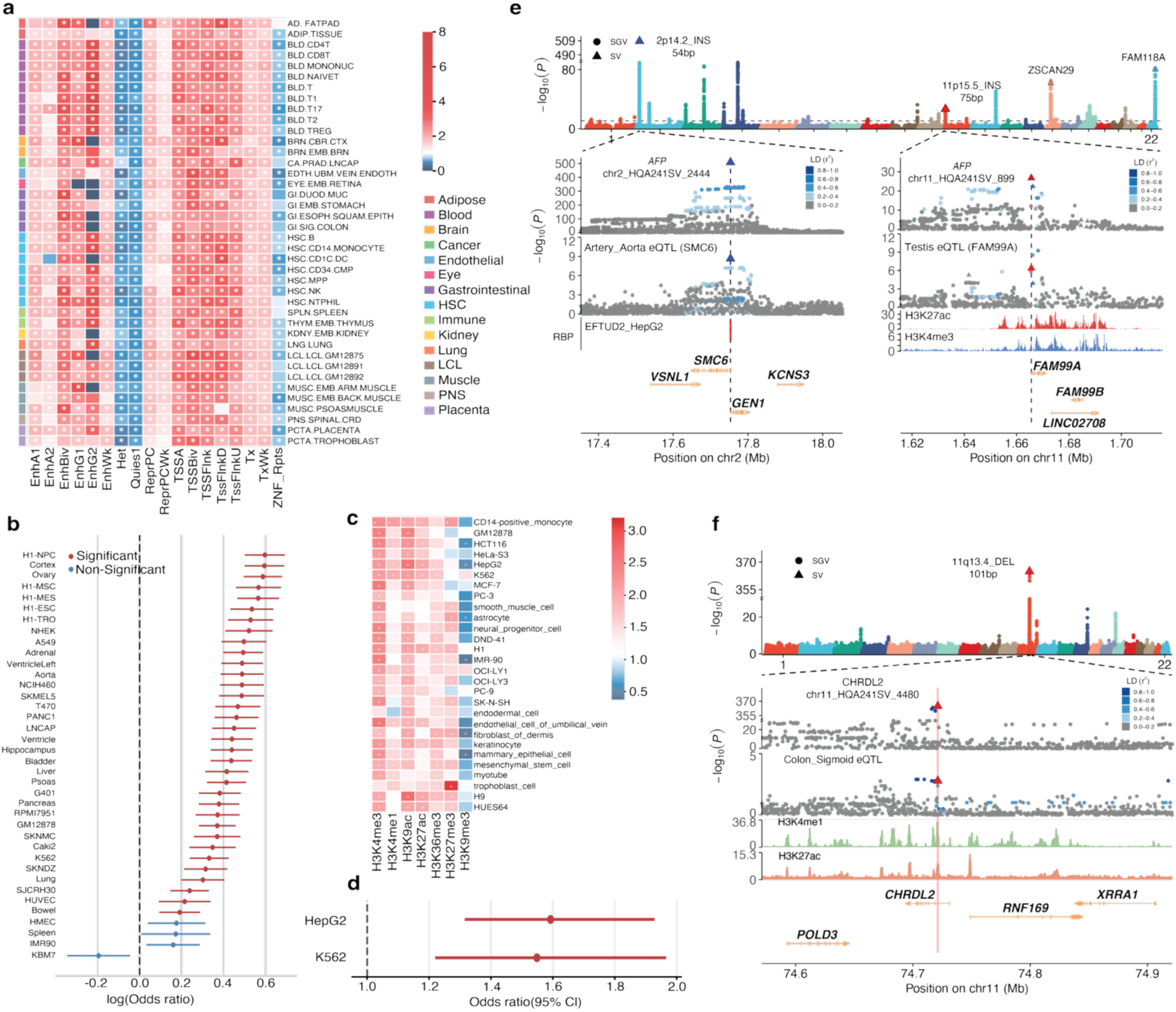
Functional enrichment analysis of SV-pQTLs. a) Enrichment of SV-pQTLs across EpiMap chromatin states. Odds ratios were estimated using logistic regression and are shown on a blue-to-red scale. Asterisks indicate FDR < 0.05 , with FDR controlled using the Benjamini–Hochberg procedure. b) Enrichment of SV-pQTLs at TAD boundaries. c) Enrichment of SV-pQTLs across histone modification marks. Odds ratios were estimated using logistic regression, and asterisks were defined in the same manner as in the chromatin-state analysis. d) Enrichment analysis results for SV-pQTLs at RBP binding sites. e) Genome-wide Manhattan plot for AFP and regional association plots for 2p14.2-INS-54bp and 11p15.5-INS-75bp, with dashed lines indicating the insertion position. Below, locus plots are shown for *SMC6* in GTEx Artery Aorta and for *FAM99A* in GTEx Testis, together with HepG2 RBP binding sites (red boxes marking RBP-bound regions). H3K27ac data are from liver (ENCFF764VSN), and H3K4me3 data are from liver (ENCFF917LFF). f) Genome-wide Manhattan plot for CHRDL2 and a regional association plot for 11q13.4-DEL-101bp, with the deleted segment highlighted in red. Below, a locus plot in GTEx Colon Sigmoid is shown, followed by tracks of H3K4me1 (ENCFF615ABM) and H3K27ac (ENCFF762YWL) in thoracic aorta.

First, we examined alpha-fetoprotein (AFP), a key hepatocellular carcinoma biomarker^29,30^. We found that circulating AFP levels are influenced by multiple remote, *trans*-acting SVs. The most significant signal was a 54-bp insertion (2p14.2-INS-54bp, chr2:17,753,324, fine-mapping PIP = 1) located near the 5’ UTR of *SMC6,* overlapping with its spliceosomal binding site^27^. Colocalization analysis using COLOC^31^ indicated that this SV-pQTL potentially shares a causal variant with an SV-eQTL for *SMC6* expression in the liver and aorta (COLOC PP. H4 = 1.0), suggesting that the insertion may disrupt *SMC6* splicing and expression. Given the role of *SMC6* in DNA repair and liver cancer, this implies that *SMC6*-mediated genomic instability may contribute to AFP dysregulation^32^. Another SV-pQTL was identified as a 75 bp insertion (11p15.5-INS-75bp, chr11:1,665,594) in the promoter of *FAM99A*, a putative tumor suppressor lncRNA^33^, with a high fine-mapping PIP of 0.98. In the liver and testis, this SV-pQTL signal colocalized with the SV-eQTL signal for *FAM99A* expression (Fig. 4e), suggesting it may modulate AFP levels through the regulation of *FAM99A*. These findings emphasize that *trans*-acting SVs in distal regulatory genes can significantly affect circulating AFP, offering new insights into the genetic regulation of this clinical biomarker.

As a second case, we highlight a deletion within a well-defined regulatory element that modulates *CHRDL2* levels. A 101-bp deletion (11q13.4-DEL-101bp, chr11:74,721,141-74,721,241), located in an intronic enhancer of *CHRDL2* (GeneHancer ID: GH11J074716), was the lead pQTL among all associated SVs and SGVs for its protein abundance. *CHRDL2* is highly expressed in gastrointestinal tissues, and this SV-pQTL signal showed strong colocalization with SV-eQTL signal for *CHRDL2* expression in colon tissue (COLOC PP. H4 = 0.98), supporting a regulatory link between the enhancer deletion and gene expression in a relevant cellular context (Fig. 4f). This finding illustrates how SVs can influence protein abundance by perturbing regulatory elements, with detectable effects in tissues where the target gene is robustly expressed.

### Integrative analysis of SV-pQTLs with SV-eQTLs and trait-associated SVs

To elucidate the potential regulatory mechanisms underlying SV-pQTLs and assess their downstream phenotypic relevance^34^, we integrated our SV-pQTL findings with SV-gene expression associations from GTEx v8^35^ and SV-trait associations in the UKB. This integrative framework links SVs to intermediate molecular phenotypes at the transcript and protein levels and connects these molecular effects to complex traits, including diseases.

We performed a comprehensive colocalization analysis using COLOC^31^ and SMR & HEIDI^36^ to identify loci at which the same SV jointly influenced circulating protein abundance and tissue-level gene expression in GTEx v8. For SV-eQTL summary statistics, we leveraged results from our previous study^5^, in which SV genotypes were imputed into 694 unrelated GTEx v8 individuals and SV-gene expression associations were mapped using OSCA^37^. We then performed colocalization across tissues for the 367 *cis*-SV-pQTL associations among the 601 SV-protein pairs prioritized above and identified 1,776 colocalized signals between SV-pQTLs and SV-eQTLs (Fig. 5d; Supplementary Table 8) across multiple tissues, meeting significance criteria of *P*_SMR_ < 1.36 × 10^−5^ (Bonferroni correction for multiple testing) with no evidence of heterogeneity in HEIDI tests ( *P*_HEIDI_ > 0.05 ) or strong colocalization support in COLOC analyses (PP. H4 > 0.8). Overall, 60.5% (222 out of 367) of these loci showed evidence of shared genetic association signals, and 171 loci exhibited shared signals across multiple tissues. These results suggest that SV-mediated regulation of gene expression contributes substantially to variation in circulating protein levels.

**Figure 5.**
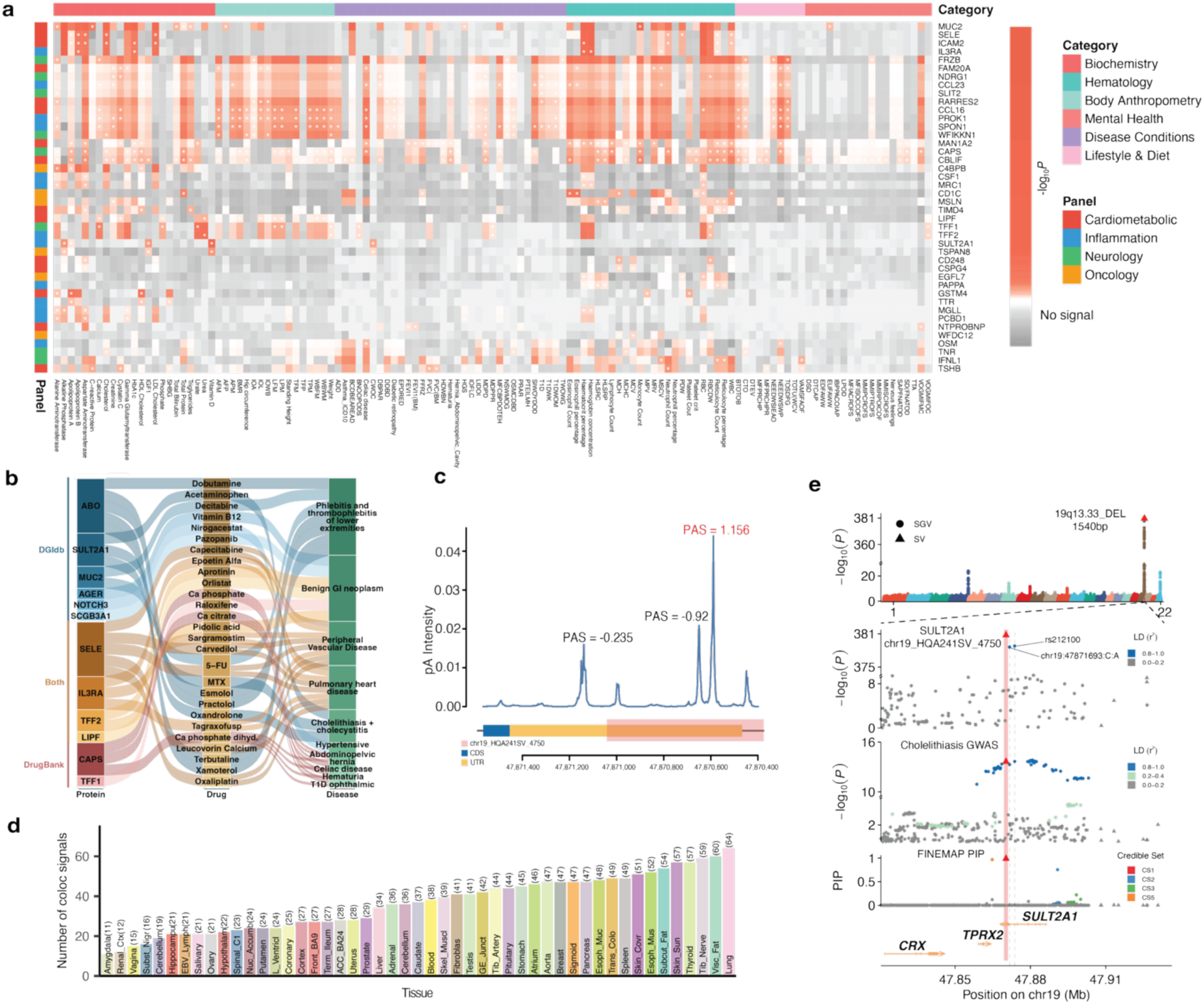
Integrative analysis of SV-pQTLs with GTEx eQTLs and UKB complex traits. a) SMR results integrating SV-pQTLs with 2,624 complex traits in the UKB. Color intensity indicates the magnitude of *P*_SMR_ , and asterisks mark associations meeting both *P*_S*MR*_ < 1.32 × 10^−5^ and the required *P*_HEIDI_ > 0.05. b) The left panel shows the drug-interacting proteins identified in our study. The middle panel shows drugs that interact with these targets, and the right panel shows the corresponding traits from the colocalization results. c) APARENT predictions across the *SULT2A1* 3’ UTR. Genomic coordinates are shown on the x-axis, with the SV highlighted in red. The y-axis indicates the APARENT-predicted cleavage and polyadenylation intensity at each position. Predicted polyadenylation signals are annotated along the 3’ UTR. d) Number of colocalized SV–pQTL and SV– eQTL signals detected in each GTEx tissue, aggregated across coloc and SMR support. Tissues are ordered by ascending signal count. e) Genome-wide Manhattan plot for SULT2A1 protein levels highlighting a 1.54-kbp deletion at 19q13.33, with regional association plots for the deletion at the SULT2A1 pQTL and for cholelithiasis with cholecystitis, together with fine-mapping posterior inclusion probabilities across the locus.

To link SV-driven changes in protein abundance to complex phenotypes, we used SMR & HEIDI^36^ to integrate SV-pQTLs identified for 2,922 proteins with SV-trait association results from our previous analysis of 2,624 traits in the UKB^5^. We focused on the colocalization between the 601 prioritized SV-protein associations and genome-wide significant SV-trait associations. Overall, 24.8% (149 of the 601) of the SV-pQTLs showed evidence of colocalization with at least one complex trait GWAS signal, yielding a total of 1,353 associated protein-trait pairs (Fig. 5a; Supplementary Table 9), meeting Bonferroni-corrected significance criteria of *P*_SMR_ < 1.32 × 10^−5^ with no evidence of heterogeneity in HEIDI tests (*P*_HEIDI_ > 0.05) or strong colocalization support in coloc analyses (PP. H4 > 0.8). A notable pleiotropic example is the 6p22.1-INS-52bp (chr6:28,044,374) SV-pQTL locus, which colocalized with GWAS signals for 86 traits in the UKB. This 52-bp insertion is located within an intronic region of the *MAN1A2* gene, which encodes an α-mannosidase involved in protein N-glycosylation, a process essential for protein folding and function^38,39^. Consistent with the established roles of N-glycosylation in cardiovascular, immune, and metabolic regulation, the 6p22.1-INS-52bp SV exhibited broad pleiotropic associations, spanning blood biochemical traits (*m* = 25), lifestyle or behavioral traits (*m* = 28), disease phenotypes (*m* = 7), and other traits (*m* = 26; Supplementary Table 10).

Here, we highlight the *SULT2A1* locus as an example from our integrative analysis, which was also implicated by Eldjarn et al.^22^ in a large-scale UKB plasma proteogenomic study linking a *cis*-SGV-pQTL for SULT2A1 to gallstone-related disease. In the Eldjarn et al. study, the prioritized variant was an SGV (chr19:47871693:C:A) in strong LD with the lead gallstone-associated SGV (rs212100). However, when SVs were included in the analysis, the signal resolved to a 1.54-kbp deletion in the *SULT2A1* 3’ UTR (19q13.33-DEL-1.54kbp, chr19:47,869,502-47,871,041). This SV, rather than the previously implicated intronic SGVs, was retained in COJO stepwise conditional analysis and assigned a PIP of 1.0 by SuSiE fine-mapping (Fig. 5e). Importantly, this deletion also maps to a functionally compelling region: APARENT^40^ predicted it to overlap the most efficient polyadenylation site in the *SULT2A1* 3’ UTR under the reference sequence context ( PAS = 1.16 ), suggesting that the SV may disrupt alternative polyadenylation, thereby altering transcript termination, mRNA stability, and ultimately protein abundance (Fig. 5c). Taken together, these findings suggest that our prioritized SV is more likely than the two previously reported intronic SGVs to be the functional variant driving the genetic association signal at this locus, with the SGVs merely tagging the effect of the SV. Consistent with this regulatory hypothesis, SMR analyses integrating SV-eQTL evidence from GTEx and SV-trait association results from the UKB showed that the SV-pQTL signal at this locus colocalized with the SV-eQTL signal for *SULT2A1* expression in the liver ( PP. H4 = 0.98 ). This SV-pQTL signal also colocalized with SV-GWAS signals for cholelithiasis with cholecystitis (*P*_SMR_ = 3.04 × 10^−14^, *P*_HEIDI_ = 0.124) and serum alkaline phosphatase (ALP) levels (*P*_SMR_ = 3.72 × 10^−14^, *P*_HEIDI_ = 0.145). Together, these findings support a plausible model in which disruption of post-transcriptional regulation by a 3’ UTR deletion links altered SULT2A1 protein abundance to hepatobiliary disease-related traits (Supplementary Fig. 3).

To evaluate the translational relevance of disease-associated SV-pQTLs, we integrated approved drug-target annotations from both the Drug-Gene Interaction Database (DGIdb)^41^ and DrugBank^42^ for known drug-gene interactions involving protein-coding genes corresponding to 33 of the disease-associated SV-pQTLs. Among these, 15 proteins (45.4%) were annotated as known drug targets, whereas 18 proteins (54.5%) had no reported drug interactions in the DGIdb and DrugBank. Of the 15 proteins with known drug interactions, twelve (ABO, AGER, CAPS, IL3RA, LIPF, MUC2, NOTCH3, SCGB3A1, SELE, SULT2A1, TFF1 and TFF2) were linked to FDA-approved drugs, with the remaining three targets (FRZB, ICAM2 and OSM) associated with investigational or experimental compounds (Fig. 5b). As an illustrative example, SULT2A1, which was associated with cholelithiasis with cholecystitis, is targeted by multiple drugs, including agents originally indicated for rheumatoid arthritis, antipyretic-analgesic therapy, and body-weight restoration-related indications. Together, these results highlight potential opportunities for drug repurposing and provide a framework for prioritizing SV-implicated proteins for therapeutic development.

To further extend our investigation into the disease relevance of SV-pQTLs, we leveraged disease-associated SGVs from the GWAS Catalog to assess whether our prioritized SVs were in high LD (*r*^2^ > 0.8) with the lead GWAS SGVs or shared the same fine-mapping credible sets at disease-associated loci^22^ (Methods). Among the 601 prioritized SV-pQTLs, 123 harbored at least one GWAS lead SGV. In total, we identified 318 pairs of SV-pQTLs and GWAS lead variants, involving 1,293 disease-relevant phenotypes (Supplementary Table 11). Given that our procedure for prioritizing SV-pQTLs explicitly accounts for SGV effects, it is highly plausible that these disease associations, previously attributed to SGVs, may instead be driven by the underlying SVs (Supplementary Table 12). While these results highlight the broad clinical relevance of SV-pQTLs, we caution that this analysis remains exploratory and will require formal validation as well-powered SV-GWAS data become available for these phenotypes.

### Genome-wide landscape of VNTR-pQTLs

We next extended our association analyses to VNTRs. Using a genome-wide significance threshold of *P*_VNTR−pQTL_ < 1.7 × 10^−11^ ( 5 × 10^−8^ ∕ 2,922 ), we identified 4,101 VNTR-protein associations (VNTR-pQTLs), including 2,252 *cis*-VNTR-pQTLs and 1,849 *trans*-VNTR-pQTLs, involving 2,075 distinct VNTRs and 1,177 proteins (Supplementary Fig. 4 and Supplementary Table 13). Leveraging VNTR-GWAS results from our previous study in the UKB^5^, we found that 2,128 (51.9%) VNTR-pQTL loci also showed genome-wide significant VNTR-GWAS evidence (Supplementary Table 14). We then applied SuSiE^25^ to fine-map VNTR-pQTL signals and obtained 1,213 VNTR-pQTL loci where the VNTR had the highest PIP (Supplementary Table 15). Similar to general SVs, VNTRs exhibited extensive polygenicity and pleiotropy. Moreover, significant VNTRs tended to lie closer to TSS ( *P* = 1.6 × 10^−470^) and SS (*P* = 2.5 × 10^−480^, two-sided Wilcoxon rank-sum test) of the cognate gene than other VNTRs (Supplementary Fig. 5).

The distribution of motif copy number variation among VNTR-pQTLs was diverse, encompassing unimodal, bimodal, and multimodal patterns, underscoring the varied effects of VNTR dosage on protein levels (Figs. 6a-c). We performed enrichment analyses of VNTR loci using the same functional annotations applied to SVs, including TAD boundaries, RBP binding sites, histone modification peaks, and chromatin-state annotations (Supplementary Figs. 6-9). Consistent with general SV-pQTLs, VNTR-pQTLs were enriched in functionally important regions and depleted in repressive or quiescent chromatin states. The following examples illustrate how VNTR length functions as a tunable regulatory element, altering the motif copy number modulates the availability of putative transcription factor binding sites and enhancer elements, leading to dosage-dependent effects on protein levels.

**Figure 6.**
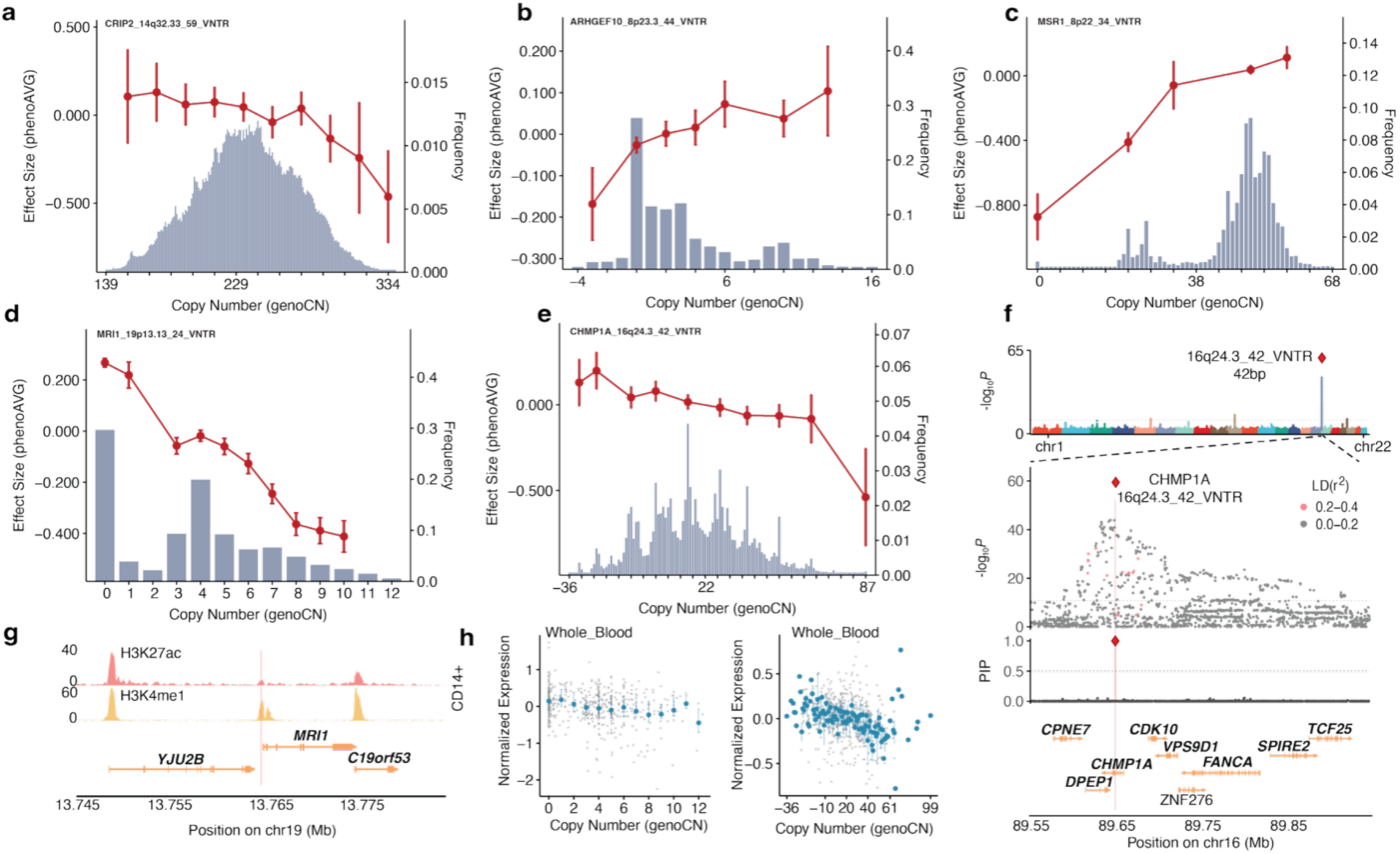
VNTR-pQTL association analysis. a–e) VNTR association results across different copy-number distributions. Blue bars indicate the frequency of each VNTR copy number, and the red line shows the mean protein abundance among individuals carrying the corresponding copy number. f) Genome-wide Manhattan plot for CHMP1A and a locus plot for 16q24.3-VNTR-42bp. The SuSiE PIPs are shown below. g) Locus tracks for 19p13.13-VNTR-51bp showing H3K27ac (ENCFF184NWF) and H3K4me1 (ENCFF587XGD) marks in CD14+ cells, with the VNTR region highlighted in red. h) Left: *MRI1* expression in GTEx whole blood as a function of 19p13.13-VNTR-51bp copy number. Right: *CHMP1A* expression in GTEx whole blood as a function of 16q24.3-VNTR-42bp copy number.

An intronic VNTR (denoted 16q24.3-VNTR-42bp), composed of a 42-bp repeat unit and located within the *CHMP1A* gene, showed a strong association between repeat length and CHMP1A protein abundance. This VNTR represented the most significant signal genome-wide at this locus, with a fine-mapping PIP of 1 (Figs. 6e,f). We compared the VNTR repeat motif against known transcription factor binding motifs using the MEME Suite^43,44^ and observed a significant sequence similarity to the *ZIC3* motif (*P* = 5.5 × 10^−5^, q-value = 0.04, significant after false discovery rate correction), suggesting that the VNTR motif resembles binding motifs of *ZIC3* or other *ZIC* family transcription factors (Supplementary Fig. 10). More generally, the presence of a repeat-length-variable motif within an intronic region raises the possibility that the VNTR influences *CHMP1A* regulation by altering local regulatory sequence properties or transcription factor recognition in a context-dependent manner. Consistent with a regulatory role for this VNTR, *CHMP1A* has been implicated in pontocerebellar and cerebellar hypoplasia^45^, and analysis of GTEx data across multiple tissues revealed VNTR-dependent changes in *CHMP1A* expression, including in whole blood (*P* = 1.1 × 10^−8^), that mirror the pattern observed in our SV-pQTL analysis (Fig. 6h).

Another VNTR (denoted 19p13.13-VNTR-51bp), composed of a 51-bp repeat unit, is located within a putative enhancer region (GeneHancer ID: GH19J013763) of the *MRI1* gene (Fig. 6d,g). Across alleles at this locus, increasing VNTR copy number was associated with progressively lower circulating MRI1 protein abundance, consistent with a dosage-dependent relationship. In our VNTR-eQTL analysis, the same VNTR showed a strong *cis* association with *MRI1* mRNA levels in GTEx whole blood (*P* = 1.6 × 10^−8^; Fig. 6h), with an effect direction concordant with that observed for the protein. Together, these findings suggest that length variation at 19p13.13-VNTR-51bp may influence enhancer activity, leading to altered *MRI1* expression in blood and, in turn, variation in plasma MRI1 protein levels.

## Discussion

In this study, we performed genome-wide association analyses of imputed SVs and VNTRs for 2,922 proteins, constructing the most comprehensive atlas to date of systematic associations between these complex SVs and circulating protein abundance. We demonstrated that SVs and VNTRs made substantial contributions to the heritability of protein abundance. By integrating functional genomic annotations, eQTL data, and GWAS results, we elucidated plausible mechanisms underlying these SV-protein associations, ranging from direct gene disruption to regulatory perturbations mediated by gene expression. Furthermore, by linking specific SV-pQTLs to complex traits, our analyses highlight promising avenues for therapeutic prioritization and drug repurposing.

Distinguishing true SV-specific effects from those merely tagging SGVs is a persistent challenge due to extensive LD. To address this, we used a stringent strategy that prioritized 601 of the 8,065 identified SV-pQTLs. These prioritized loci represent distinct associations highly unlikely to be driven by linked SGVs, as they satisfy at least one of the following criteria: acting as the lead GWAS variant, retaining genome-wide significance after conditioning on correlated SGVs, or emerging as the primary candidate in SuSiE fine-mapping analyses. However, because SV genotyping accuracy is generally lower than that of SGVs, our statistical power to definitively uncouple SV effects from linked SGVs remains constrained. Consequently, the SV-pQTLs not prioritized in this analysis likely contain many true structural regulatory effects, making them valuable targets for future in-depth investigation.

Our colocalization analyses, integrating SV-pQTLs with gene expression associations, provided critical insights into the mechanistic roles of SVs in biological processes. A notable example involves AFP, a widely used tumor biomarker in clinical practice. Our analyses revealed that multiple distal SVs contribute to variation in circulating AFP levels. This includes a *trans*-acting SV-pQTL near the 5’ UTR of *SMC6* that colocalizes with its *cis*-acting SV-eQTL, suggesting a potential role in regulating AFP levels through effects on *SMC6*, a gene implicated in DNA repair, RNA processing, and cancer pathways^29,30,32,33^. These observations suggest that SV-mediated perturbation of regulatory networks can affect AFP protein abundance via *trans*-regulatory mechanisms, highlighting an additional layer of genetic control beyond variants located near the *AFP* locus itself. More broadly, these findings illustrate how distal SVs influence clinically important biomarkers through diverse mechanisms, providing new insights into their genetic regulation. Furthermore, integrating SV-pQTLs with SV-GWAS for complex traits revealed 1,353 protein-trait links underpinned by shared SV signals. This demonstrates that many SV-pQTLs colocalize with established disease loci, pointing to shared structural genetic determinants and nominating these proteins as plausible mediators of disease risk. These genetically supported protein-disease links provide a framework for therapeutic prioritization by highlighting SV-driven alterations in circulating proteins, which serve as measurable readouts of underlying disease mechanisms potentially amenable to pharmacological intervention. For example, the SV-pQTL signal for MAN1A2 protein abundance, driven by the 6p22.1-INS-52bp insertion, colocalized with GWAS signals for 86 traits. This extensive pleiotropy aligns with the central role of MAN1A2 in protein N-glycosylation, reflecting the broad physiological impact of perturbing a fundamental protein maturation pathway.

Our study also highlights the broad disease relevance of SV-pQTLs and suggests that a subset of previously reported disease-associated SGV-pQTLs may in fact be attributable to underlying SVs. For example, a shared SV association signal linked *SULT2A1* to cholelithiasis with cholecystitis, implicating this gene in gallbladder-related disease risk. Given the role of SULT2A1 in hepatic steroid and bile acid metabolism, its altered regulation may influence biliary composition and inflammatory pathways relevant to gallstone disease. Notably, multiple drug-gene interactions reported in the DGIdb and DrugBank indicate that several SV-pQTL-implicated proteins, including SULT2A1, are already pharmacologically tractable. These observations highlight opportunities to leverage SV-informed pQTL mapping for prioritizing therapeutic targets and identifying candidates for drug repurposing by connecting existing pharmacological agents to genetically supported disease mechanisms. Ultimately, our SV-pQTL resource provides a robust framework for linking structural genetic variation to disease-associated proteins, guiding future therapeutic discovery.

We contextualized our findings alongside recent UKB SV-pQTL studies utilizing short-read WGS^11^ and moderate-coverage ONT imputation^4^. Although we observed substantial consistency, recovering 61.1% and 35.1% of the prioritized signals reported in the two studies, respectively (Supplementary Tables 16 and 17), direct quantitative comparisons were constrained by the unavailability of complete summary statistics, differing significance thresholds, and distinct variant prioritization frameworks across studies. Instead, study-specific associations highlight the complementary nature of different SV detection strategies. Our approach relies on the unparalleled sequencing accuracy of a haplotype-resolved, long-read assembly-based reference panel. This design successfully resolves complex, low-mappability, and repetitive regions where short-read and moderate-coverage technologies struggle with precise breakpoint definition and genotyping accuracy. By overcoming these limitations, our framework exclusively identified 494 of our 601 prioritized SV-pQTLs (Supplementary Table 18), demonstrating the critical importance of reference panel accuracy and substantially expanding the known structural regulatory landscape.

Despite these advances, our study has several limitations. First, because SV and VNTR genotypes were obtained through imputation, the incomplete capture of rare variants and imperfect imputation accuracy may lead us to underestimate the effects of SVs on protein levels. This limitation should diminish as long-read sequencing cohorts expand to population scale and as SV detection technologies continue to improve, mirroring the historical trajectory of SGV discovery. Second, our study was restricted to individuals of European ancestry from the UKB, primarily due to sample size constraints; larger, multi-ancestry cohorts will be required to discover additional signals. Third, all protein abundances were quantified from circulating plasma. Although some tissue-specific proteins can be detected in plasma, measurements from a broader range of tissues will be needed to better capture context-specific regulation. Finally, we cannot completely rule out epitope-binding artifacts, because data from orthogonal platforms were unavailable for validation; nevertheless, by assessing LD with PAVs and leveraging eQTL evidence, we aimed to minimize the inclusion of loci likely influenced by epitope effects.

In summary, our comprehensive genome-wide analysis of circulating plasma proteins reveals widespread associations with SVs and VNTRs, highlighting their diverse functional implications. These results establish a key resource for probing the molecular mechanisms of SV-pQTLs and investigating the contribution of SVs and VNTRs to complex traits and diseases.

## Online Methods

### Proteomics data in the UK Biobank

Protein abundance data were obtained from the UKB Pharma Proteomics Project (UKB-PPP), which profiled circulating plasma proteins in UKB participants using the Olink platform. The dataset comprised measurements of 2,923 plasma proteins across 54,219 participants and was generated using antibody-based assays designed to quantify a broad range of biologically and clinically relevant proteins. Proteomic data underwent standard quality control procedures as described in the original study^9^. Following the original report, we excluded GLIPR1 because approximately 80% of its measurements failed quality control, resulting in a final set of 2,922 proteins for downstream analyses^9^. Protein measurements were further processed by removing extreme values exceeding five standard deviations from the mean, followed by a rank-based inverse normal transformation to normalize the distributions.

### SV and VNTR imputation

SVs and VNTRs were imputed into the UKB dataset using the same pipeline and reference panel as described in our previous work^5^. Briefly, imputation was performed on the UKB Research Analysis Platform using TOPMed pre-imputed SNP genotypes generated by the UKB team. SV and VNTR imputation was conducted using a haplotype-resolved, long-read assembly-based reference panel, with imputation carried out using Minimac4 (v4.1.6)^46^. In the present study, we restricted analyses to UKB participants of European ancestry. Ancestry assignments were based on principal component projection onto the 1000 Genomes Phase 3 reference panel^47^, and individuals who had withdrawn consent were excluded. A total of 54,219 unrelated individuals of European ancestry were included in downstream analyses. Post-imputation quality control followed the same criteria as in our previous study. Specifically, imputed SVs were retained if they had a MAF ≥ 0.01, an imputation INFO score ≥ 0.3, and did not deviate from HWE (*P* > 1 × 10^−6^). VNTR loci with poor imputation quality, defined by a *r*^2^ < 0.1 in benchmarking analyses, were excluded. After filtering, the final dataset comprised 54,578 common SVs and 15,826 VNTRs in the European-ancestry subset, which were used for all subsequent analyses.

### Variant-based heritability estimation of plasma proteins

We estimated the variant-based heritability of 2,922 proteins attributable to SGVs, SVs, and VNTRs using GCTA-GREML^18,48^. For SVs and SGVs, we used GCTA^17^ to compute genetic relationship matrices (GRMs) based on variants from 46,774 unrelated individuals. For VNTRs, we employed dgrm (https://github.com/PeixiongYuan/dgrm) to estimate the genetic relationship between individuals *j* and *k* using the following formula: 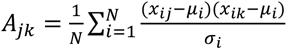, where N is the number of VNTRs, *x* and *x_ik_* are the genotypes (dosage values) of individuals *j* and *k* at VNTR *i*, and *μ_i_* and *σ_i_* are the empirically estimated mean and standard deviation of genotypes at VNTR *i* . This approach is consistent with the GRM calculation implemented in GCTA but uses empirical variance rather than allele-frequency-based theoretical variance, which is more suitable for VNTR dosage data. We first estimated heritability separately for SGVs, SVs, and VNTRs, and then fitted the three corresponding GRMs jointly to partition the heritability. The following covariates were included in all models: age, age squared, sex, age × sex, age squared × sex, the first 20 genetic principal components (PCs), time course, eight assay batches, and 22 UK Biobank assessment centers. To avoid selection bias, we applied the --reml-no-constrain option. To quantify the variance explained by individual variants, we calculated per-variant heritability using the fastGWA^21^ effect estimates and the formula: 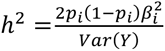, where *p_i_* is the allele frequency of variant *i* in the analysis sample, *β_i_* is the fastGWA-estimated additive effect size for variant *i*, and *Var*(*Y*) denotes the phenotypic variance of the trait after covariate adjustment.

### Genome-wide association analyses of imputed SVs and VNTRs for plasma proteins

We performed genome-wide association analyses of 54,578 imputed SVs and 8,801,428 TOPMed-imputed SGVs for the abundances of 2,922 plasma proteins. Association testing was conducted using GCTA-fastGWA^21^. To account for sample relatedness, we constructed a sparse GRM based on SGVs. The covariates included in these analyses were identical to those used in the GREML heritability analyses. For VNTR association analyses, to control for sample relatedness and population structure while maximizing sample size and statistical power, we first applied GCTA-fastGWA using the same covariates as in the SV association analyses and enabled the --save-fastGWA-mlm-residual option to obtain residuals for each plasma protein measurement. We then performed linear regression of these protein residuals on VNTR dosage in R.

### Sensitivity analyses of SV-pQTLs

To minimize post hoc bias, we performed sensitivity analyses of the associations between SVs and plasma protein levels, additionally extracting blood cell counts, blood collection time, and body mass index (BMI) from the UKB. For blood cell traits, we extracted data on monocyte count, basophil count, lymphocyte count, neutrophil count, eosinophil count, leukocyte count, platelet count, haematocrit percentage, haemoglobin concentration, and mean corpuscular volume. We removed extreme outliers, excluding samples with a leukocyte count > 200 × 10^9^ ∕ L or (a leukocyte count > 100 × 10^9^ ∕ L and immature reticulocytes > 5% ), haemoglobin concentration > 20g/dL , haematocrit > 60%, or platelet count > 1000 × 10^9^ ∕ L. For blood collection time, we obtained the season of blood draw and fasting duration. We coded winter/spring (December-May) as 0 and summer/autumn (June-November) as 1, and standardized fasting time using a z-score transformation. We then merged each of these three data types with the covariates used in the primary association analyses and re-ran the association analyses using fastGWA^21^.

### Conditional and fine-mapping analyses of SV-pQTLs

We performed conditional analyses of SV-pQTLs using GCTA-COJO^24^ to identify SV associations that remained significant after conditioning on SGVs. We used GCTA to compute a GRM and selected 46,774 unrelated individuals with a GRM cutoff of 0.05 as the LD reference panel. Conditional analyses were then conducted using a 10 Mb window and a significance threshold of *P* < 1.7 × 10^−8^. Subsequently, we applied sum of single-effects regression (SuSiE, v0.14.2)^25^ to fine-map SV-pQTL signals. For each conditionally independent SV-pQTL, we performed fine-mapping within a 1 Mb window around the lead SV. LD matrices for each region were computed using PLINK2 (v2.0.0)^23^ based on all unrelated individuals. Within each region, SVs that ranked among the variants with the highest PIPs were defined as top-PIP SVs.

### Identification of lead variants of SV-pQTLs

We used PLINK2 (v2.0.0)^23^ to perform LD-based clumping to identify approximately independent association signals. For each chromosome, we applied an LD reference panel constructed from unrelated European-ancestry individuals in the UK Biobank cohort. Clumping parameters were set as follows: genome-wide significance threshold *P* < 5 × 10^−8^, secondary significance threshold *P* < 5 × 10^−8^, LD *r*^2^ = 0.01 with a window size of 1 Mb.

### Quality control for epitope assay-binding effects

We annotated SGVs and SVs using Variant Effect Predictor (VEP, v113.0)^49^ and AnnotSV^50^, respectively. For variants located within the coding sequence or splice sites of the protein-coding gene corresponding to each SV-pQTL, we computed LD between the SV and PAVs using PLINK2 (v2.0.0)^23^. Leveraging SV-eQTL results from GTEx, we defined loci as having a higher likelihood of being affected by epitope-binding artifacts if (i) the SV was in LD with a PAV (*r*^2^ > 0.1) and (ii) there was no evidence of colocalization with an eQTL signal.

### Functional enrichment analyses of SV-pQTLs

We performed functional enrichment analyses of significant SV association loci with chromatin states, TAD boundaries, histone modifications, and RBP binding sites. For chromatin states, we obtained 18-state Roadmap model annotations from EpiMap^28^ for 11 organ or tissue groups: blood, bone, brain, eye, kidney, liver, lung, muscle, pancreas, reproductive, and spleen. For TAD boundaries, we downloaded TAD coordinates for 38 tissues or cell types from the 3D Genome Browser^26^, defined TAD boundaries as flanking regions extending ±50% of each TAD’s length, and excluded regions overlapping heterochromatin or centromeres. We obtained ChIP-seq data for key histone modifications (H3K4me1, H3K4me3, H3K36me3, H3K27me3, H3K9me3, H3K27ac, and H3K9ac) across 26 cell types from the ENCODE^27^ data portal. Low-confidence peaks with a fold enrichment of peak calling lower than 2 or *P* > 1 × 10^−5^ were removed, and the remaining peaks were sorted and merged using BEDtools (v2.29.1)^51^. For RBP binding sites, we downloaded eCLIP datasets for 140 target RBPs in K562 and HepG2 cells from ENCODE^27^, originally generated by the Gene Yeo lab. We performed enrichment analyses using logistic regression in R to compare SVs with significant association signals against non-significant SVs. To obtain independent SVs, we applied LD pruning in PLINK2 (v2.0.0) to 8,513 significant SVs and 46,065 non-significant SVs, using a 500 kb window and an LD threshold of *r*^2^ = 0.1. For each functional annotation, the logistic regression outcome indicated whether an SV overlapped the annotation, and we included SV minor allele frequency, length, variant type (insertion or deletion), and distances to the nearest TSS and SS as covariates. For each functional category, we applied FDR correction to the resulting *P* values and defined enrichment as significant at *P* < 0.05.

### Colocalization analyses

We imputed SVs into the GTEx v8 dataset as described in our previous work^5^. Briefly, SNP genotypes derived from short-read sequencing in GTEx were first imputed across 49 tissues, after which variants with a MAF < 0.01 , INFO < 0.3 , or HWE test *P* < 1 × 10^−6^ were excluded. We then performed SV-eQTL mapping using OSCA (v0.46.1)^52^ based on normalized gene expression data and covariate matrices obtained from the GTEx Portal, with gene annotations derived from the GENCODE reference. Colocalization analyses between SV-eQTL and SV-pQTL signals were performed using the coloc.abf function in the COLOC package (v5.2.3)^31^ and SMR & HEIDI (v1.3.1)^36^. Analyses were conducted within ±1 Mb windows centered on each of the 601 SV-pQTL loci prioritized as likely causal, using default parameter settings. Loci were considered colocalized when the posterior probability for a shared causal variant exceeded 0.8 (PP. H4 > 0.8) or *P*_SMR_ < 1.36 × 10^−5^ with no evidence of heterogeneity in HEIDI tests ( *P*_HEIDI_ > 0.05 ). Across the 1,776 colocalized signals identified in different tissues, we further assessed whether the effects of SVs on gene expression and protein abundance were concordant in direction.

To connect SV-pQTLs with complex traits, we leveraged our previous association analyses of 2,624 UKB traits and performed colocalization using SMR & HEIDI (v1.3.1)^36^ and COLOC (v5.2.3)^31^. For each of the 601 likely causal SV-pQTL loci, we tested colocalization with trait associations sharing the same SV within a ±1 Mb window. Within the SMR & HEIDI framework, we applied a significance threshold of *P*_SMR_ < 1.32 × 10^−5^ and required no evidence of heterogeneity with *P*_HEIDI_ > 0.05. In addition, we considered coloc results to provide strong support for colocalization when PP. H4 > 0.8.

### GWAS Catalog disease-related analysis

To assess the disease relevance of prioritized SV-pQTLs, we cross-referenced them with disease-associated variants from the NHGRI-EBI GWAS Catalog using two complementary approaches adapted from Eldjarn et al.^22^. For each prioritized SV, we computed LD with GWAS Catalog SNPs within a 1 Mb window using PLINK2 (--r2-unphased) in unrelated European-ancestry UKB individuals. SV– GWAS SNP pairs with *r*^2^ > 0.8 were considered to share a disease-associated signal. For SVs assigned to a fine-mapping 95% credible set, we further identified disease associations where a variant within the same credible set was both in high LD (*r*^2^ > 0.8) with the SV and annotated as a GWAS Catalog disease-associated variant. Associations mapped to protein measurement phenotypes were excluded from both methods.

## Author contributions

JY conceived and directed the study. PY, WB and JY designed the experiments and developed the analytical pipelines. PY, WB and JH developed the online tool and the data portal under the guidance of JY. PY performed the data analyses with assistance and guidance from WB and JY. PY, WB and JY wrote the manuscript. All authors reviewed and approved the final manuscript.

## Acknowledgements

We thank Zhongqu Duan, Shuli Liu, Yazhou Guo, Yifei Wang, Ting Xu, Xiwei Sun, Ruilei Ma and Ting Qi for helpful discussions. This research was partly supported by the National Natural Science Foundation of China (U23A20165 and 32500475), the National Key R&D Program of China (2024YFC3405800 and 2024YFC3405802), the “Pioneer & Leading Goose” R&D Program (2024SSYS0032 and 2025C01092), and the New Cornerstone Science Foundation. This research has been conducted using the UK Biobank Resource under Application Number 66982. We thank the Westlake University High-Performance Computing Center for computing support.

## Data availability

The individual-level genotype and phenotype data are available upon formal application to the UKB (http://www.ukbiobank.ac.uk). The SV and VNTR imputation panels are available at (https://yanglab.westlake.edu.cn/impute_sv). The SV- and VNTR-GWAS summary statistics generated in this study are available at (https://yanglab.westlake.edu.cn/data/sv-pqtl). The GTEx (v8) data are available at (https://www.gtexportal.org/home/datasets). The histone ChIP-seq peak and RBP eCLIP peak data are available through the ENCODE portal (https://www.encodeproject.org). The 18-state Roadmap model annotation data are available at EpiMap (https://compbio.mit.edu/epimap/). The TAD coordinate data are available through the 3D Genome Browser (https://3dgenome.fsm.northwestern.edu/datasets).

