## Supplementary Figures for "A proteome atlas of structural variation and VNTR effects on complex traits and diseases"

Yuan *et al*.

**Contents**

**Supplementary Figures 1-10**

**Supplementary References**

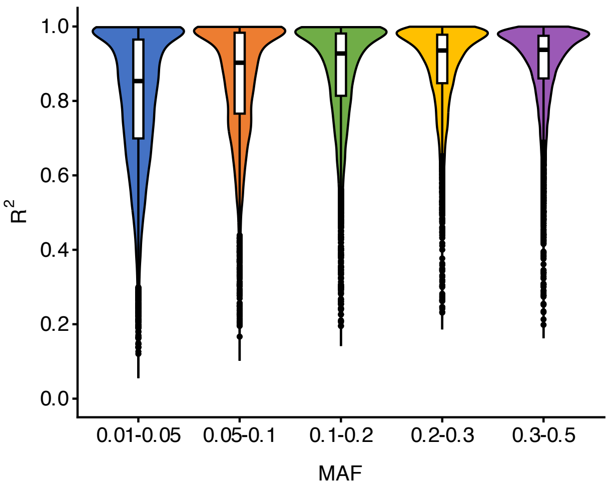


**Supplementary Figure 1. Imputation accuracy of SVs in UKB participants of European ancestry.** Violin plots show the distribution of imputation $R^2$ for 54,578 imputed SVs in European-ancestry individuals, stratified by minor allele frequency (MAF)[1]. Each color represents a distinct MAF bin. The embedded boxplots denote the median and interquartile ranges. The mean $R^2$ was 0.86, and 72.1% of SVs achieved $R^2 \geq 0.8$.

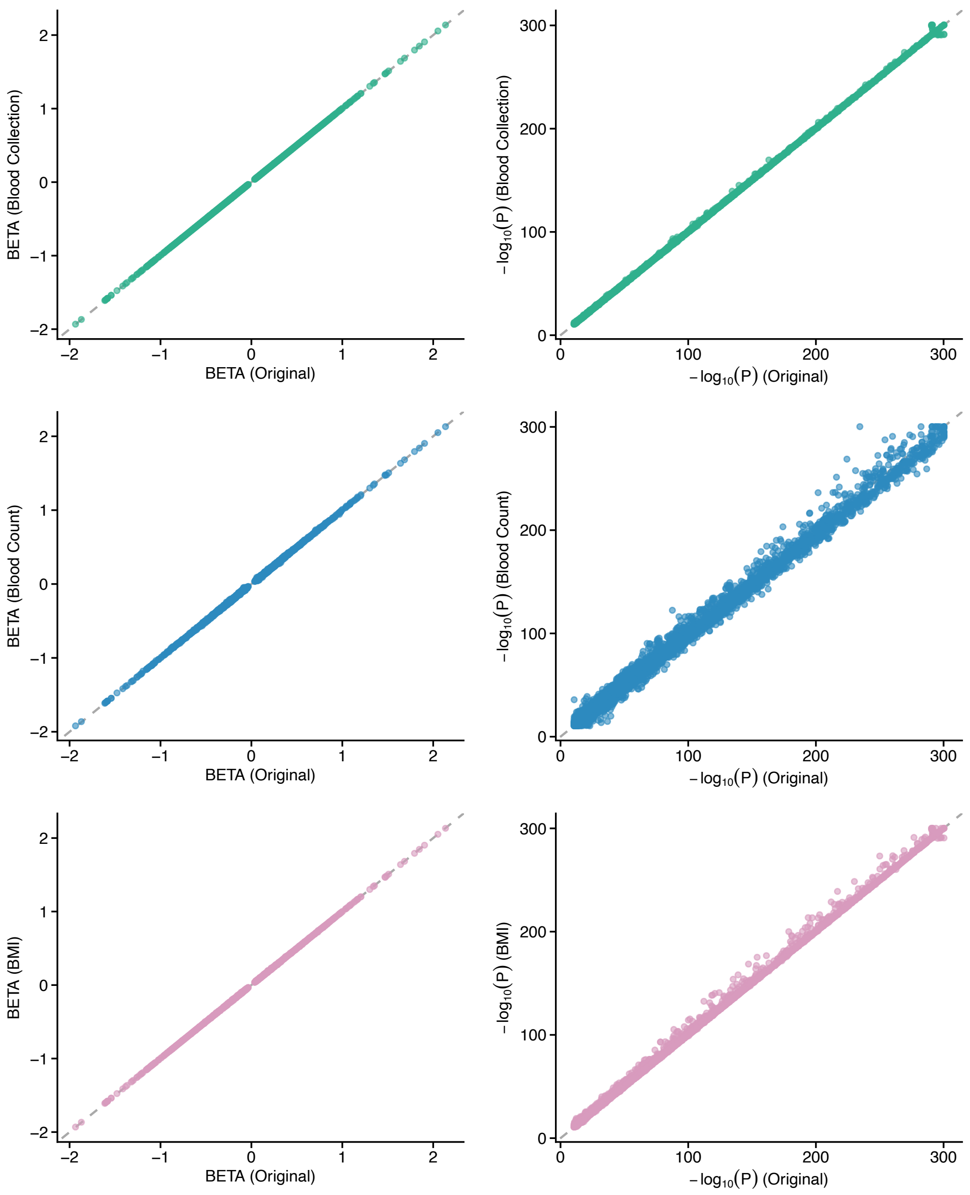


**Supplementary Figure 2. Sensitivity analysis of SV-pQTLs.** Association analyses were repeated with further adjustment for blood collection time (a), blood cell counts (b), and body mass index (BMI; c), three factors known to influence plasma protein measurements. For each covariate, effect size estimates (left panels; BETA) and statistical significance (right panels; $-\log_{10}(P)$) from the adjusted models are plotted against those from the primary analyses. The strong concordance of effect sizes and $P$ values shows that the identified SV-pQTL signals are highly robust to these potential confounders.

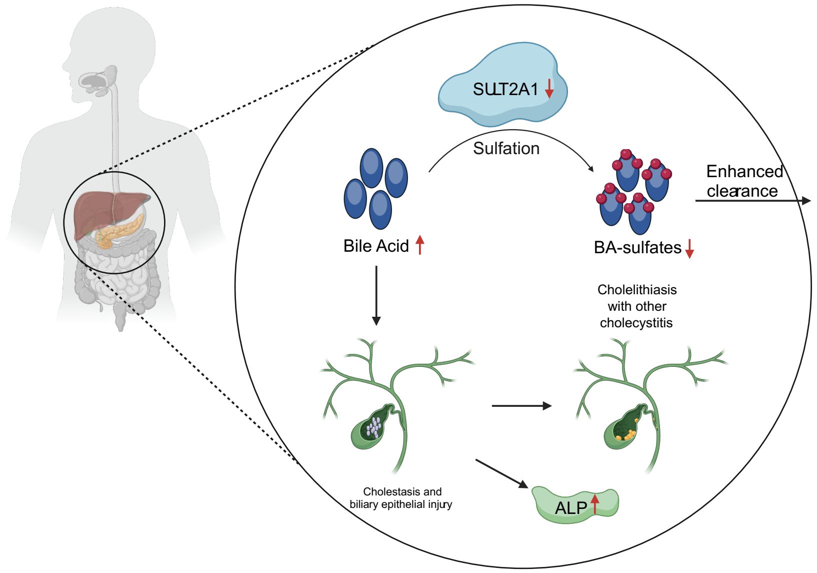


**Supplementary Figure 3. Proposed mechanism linking a 3′ UTR deletion at SULT2A1 to hepatobiliary disease traits.** A 1.54-kb deletion at 19q13.33 within the *SULT2A1* 3′ UTR is predicted to disrupt post-transcriptional regulation by overlapping the highest-efficiency alternative polyadenylation (APA) site in the reference context (APARENT[2]; PAS = 1.16), potentially altering transcriptional termination and mRNA stability and thereby reducing SULT2A1 protein abundance. Given that SULT2A1 is a bile-acid sulfotransferase, decreased SULT2A1 activity may reduce bile-acid sulfation, impair clearance, and promote bile acid accumulation, contributing to cholestasis and gallbladder inflammation[3,4]. Consistent with this model, the locus colocalizes with *SULT2A1* liver expression (GTEx[5]; $\mathrm{PP.H4} = 0.98$) and with cholelithiasis with cholecystitis and serum alkaline phosphatase (ALP) levels in the UKB (SMR & HEIDI[6]), supporting a regulatory link between the SV, altered bile-acid handling, and hepatobiliary disease–related phenotypes.

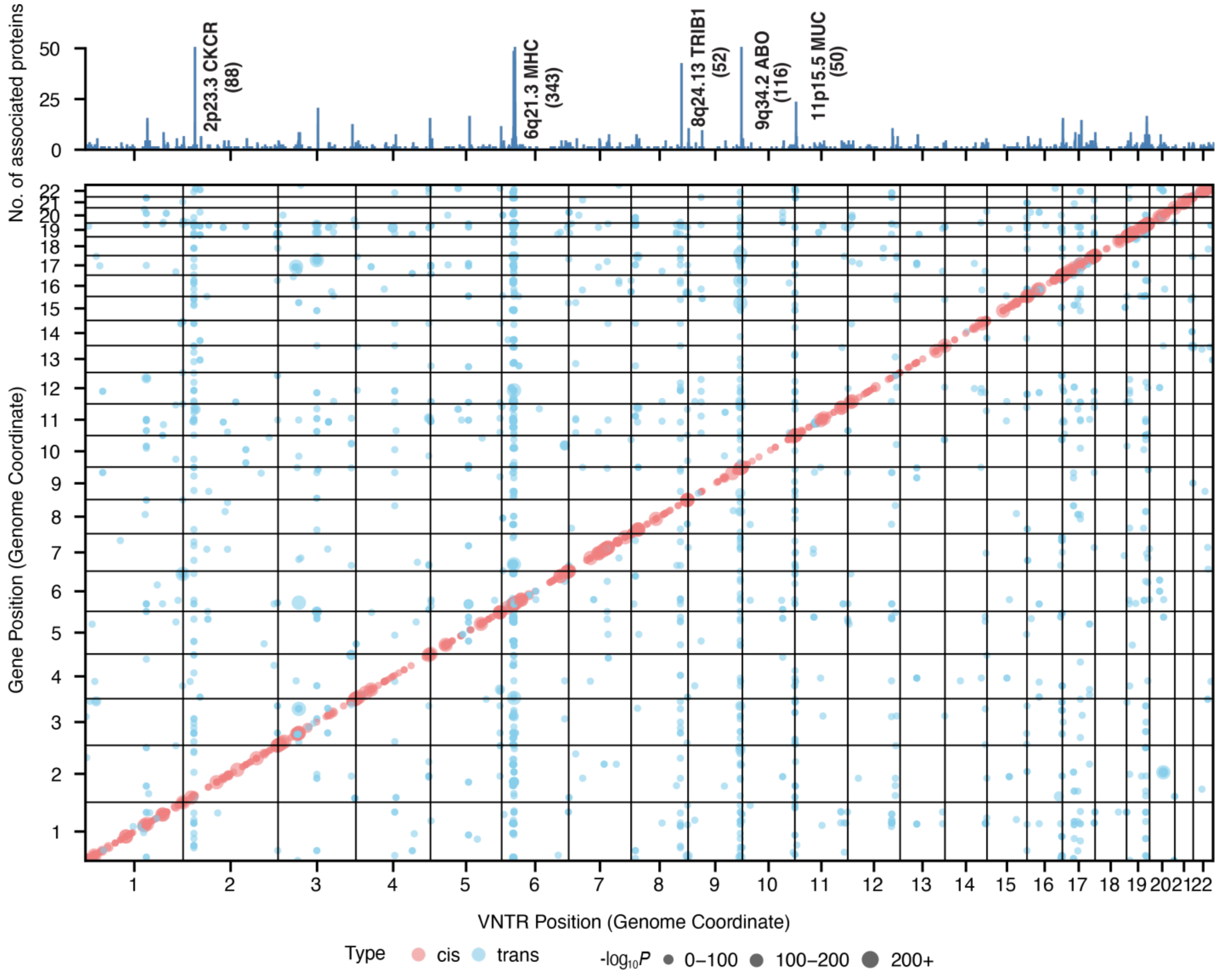


**Supplementary Figure 4. Genome-wide VNTR–protein association landscape.** The lower panel shows the genome-wide distribution of significant VNTR–protein associations, with VNTR genomic position on the x-axis and the genomic position of the corresponding protein-coding gene on the y-axis. Each point represents a significant VNTR-pQTL; colors denote *cis* (red) and *trans* (blue) effects. The upper panel summarizes the number of proteins showing significant associations in each genomic region and highlights several hotspot loci and their associated protein counts.

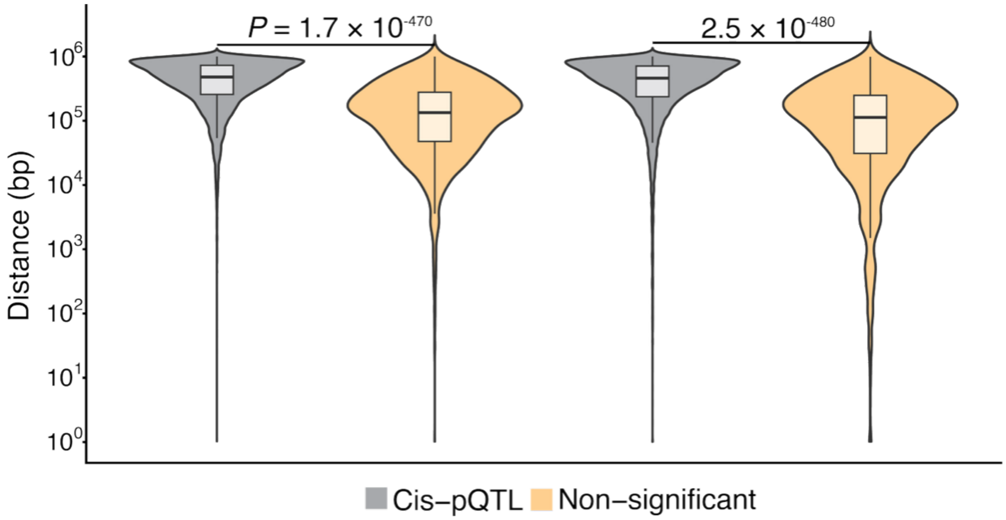


**Supplementary Figure 5. Genomic proximity of protein-associated VNTRs to regulatory landmarks.** Violin plots show the distribution of distances (bp; log scale) from VNTR breakpoints to the nearest transcription start site (TSS) (left) and splice site (right) for *cis*-pQTL VNTRs (gray) compared with non-significant VNTRs within the same *cis* window (TSS ± 1 Mb) (light orange). Embedded boxplots indicate median and interquartile range. P values (two-sided Wilcoxon rank-sum test) are shown above each comparison.

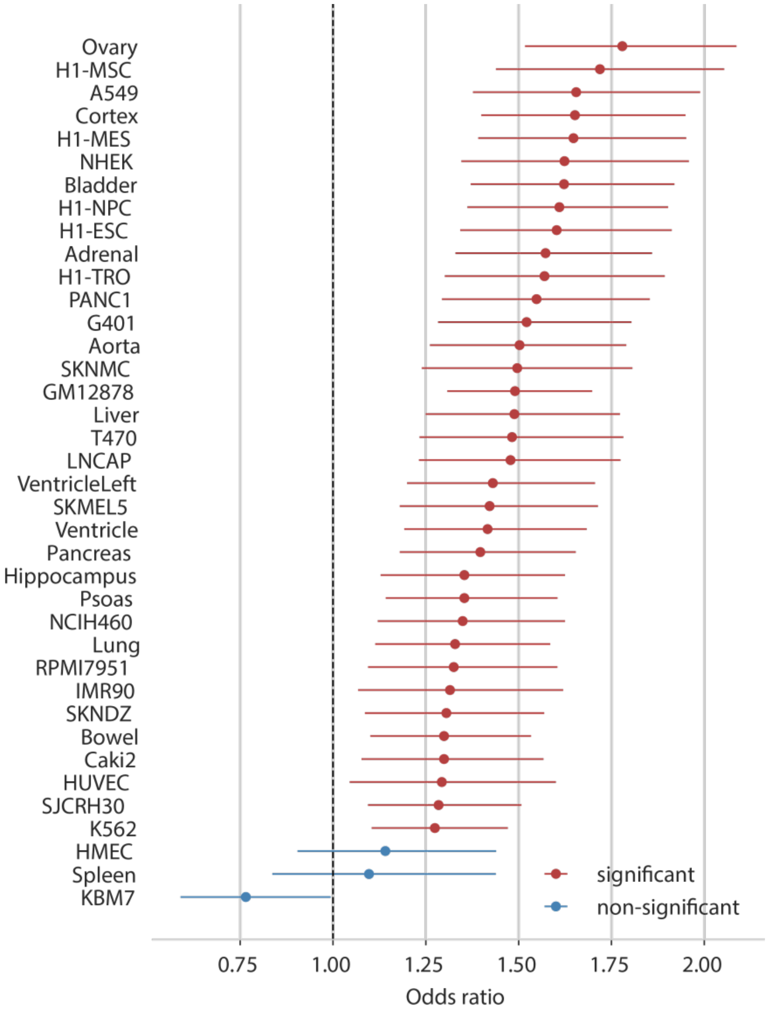


**Supplementary Figure 6. Enrichment of VNTR-pQTLs at TAD boundaries.** Forest plot showing the odds ratios (points) and 95% confidence intervals (horizontal lines) for the overlap of VNTR-pQTLs with topologically associating domain (TAD) boundaries in each cell type[7]. Enrichment was assessed by comparing genome-wide significant VNTR-pQTLs to matched background VNTRs (see Methods). The dashed vertical line indicates no enrichment (odds ratio = 1). Red points denote significant enrichment after multiple-testing correction, whereas blue points indicate non-significant results.

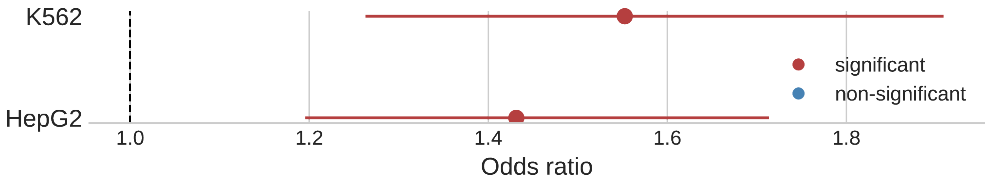


**Supplementary Figure 7. Enrichment analysis results for VNTR-pQTLs at RBP binding sites.** Forest plot showing odds ratios (points) and 95% confidence intervals (horizontal lines) for the overlap of genome-wide significant VNTR-pQTLs with RBP binding sites across cell types (K562 and HepG2)[8]. Enrichment was evaluated relative to matched background VNTRs (see Methods). The dashed vertical line indicates no enrichment ( $\text{odds ratio} = 1$ ). Red points denote significant enrichments after multiple-testing correction, whereas blue points indicate non-significant results.

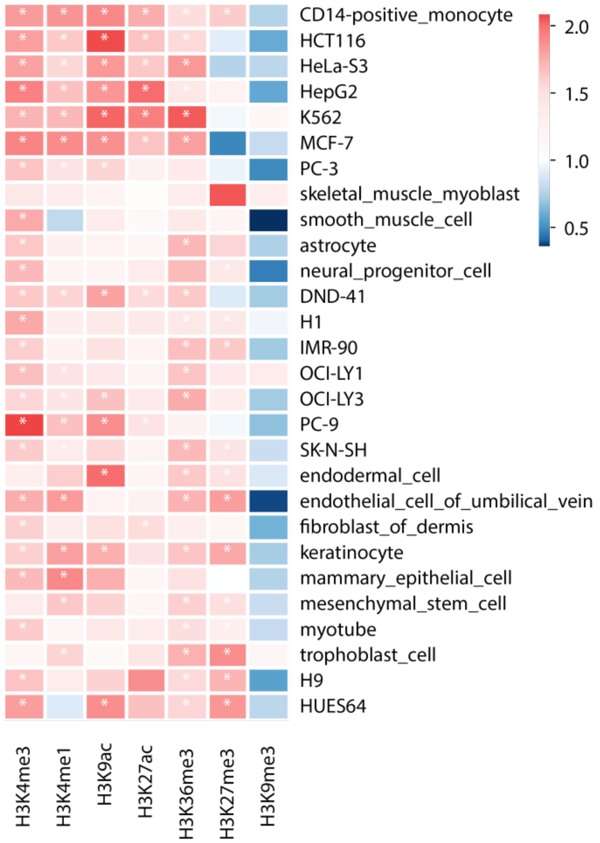


**Supplementary Figure 8. Enrichment of VNTR-pQTLs across histone modification marks.** Heatmap showing odds ratios for the enrichment of genome-wide significant VNTR-pQTLs within peaks of histone modification marks (columns) across cell types (rows)[8]. Odds ratios were estimated using logistic regression relative to matched background VNTRs (see Methods); values >1 indicate enrichment, and values <1 indicate depletion (color scale). Asterisks denote statistically significant enrichments after multiple-testing correction, defined in the same manner as in the chromatin-state analysis.

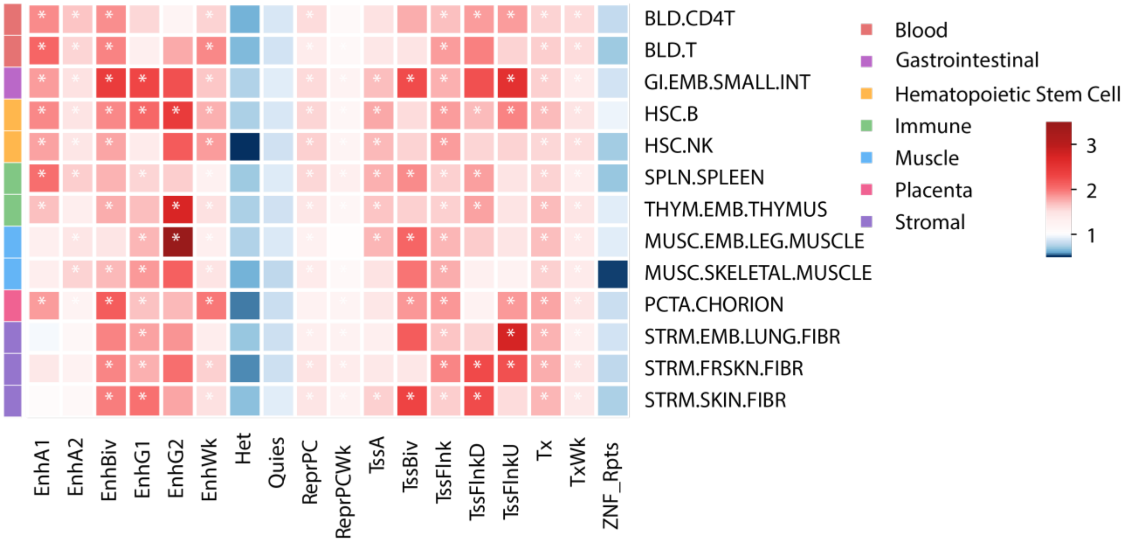


**Supplementary Figure 9. Enrichment of VNTR-pQTLs across EpiMap chromatin states.** Heatmap showing odds ratios for the enrichment of genome-wide significant VNTR-pQTLs within EpiMap chromatin states (columns) across tissues/cell types (rows; grouped by tissue category)[9]. Odds ratios were estimated using logistic regression relative to matched background VNTRs (see Methods) and are displayed on a blue-to-red scale (red, enrichment; blue, depletion). Asterisks denote significant enrichments at FDR < 0.05, with FDR controlled using the Benjamini–Hochberg procedure.

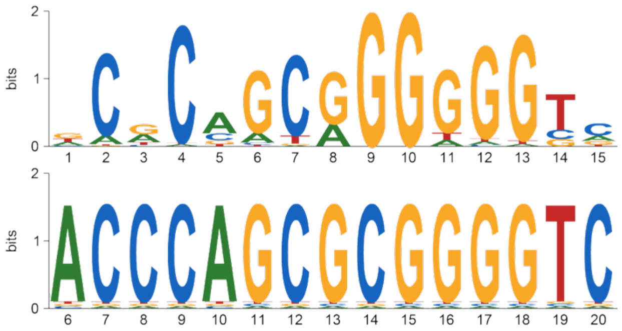


**Supplementary Figure 10. Matching the VNTR repeat motif to known transcription factor binding motifs.** The VNTR repeat consensus sequence was compared against curated transcription factor binding motifs using the MEME Suite (TOMTOM[10,11]). The top match showed significant sequence similarity to the *ZIC3* motif (Jolma2013; reverse-complement orientation), with an alignment $P = 5.5 \times 10^{-5}$ and q-value = 0.04. Sequence logos depict the reference *ZIC3* motif (top) and the aligned VNTR repeat motif (bottom); the shared high-GC core indicates that the VNTR repeat resembles binding motifs of *ZIC3* or other ZIC-family transcription factors.
